# Implementing a Behaviour Change Communication interaction for enhancing male involvement in maternity care among the Saharia Tribes in Gwalior District, Madhya Pradesh: a feasibility study

**DOI:** 10.1101/2024.07.05.24309998

**Authors:** Tulsi Adhikari, Saritha Nair, Ashpinder kaur Grover

## Abstract

**Background:** The Indian tribal population is diverse, with a wide range of customs, ways of life, and cultural practices. However, there is one thing that all Indian tribal communities have in common: they have worse health indicators, a higher rate of illness and mortality, and very restricted access to medical care. Their health issues require extra consideration in the right setting.[1] Growth in the utilization of reproductive and maternal health services will not only curtail down the reproductive morbidities, but it will also reduce the child mortality.[2] Men’s participation in prenatal care, delivery and postpartum period is rarely found, especially among tribal communities, due to their economic instabilities and priorities. Also our health system does not promote the involvement of men in the maternal and child health care. Hence, there was a need felt for development of gender and community sensitive interventions package that could address the individual and the community health care facility level barriers of male involvement in utilisation of the maternal care services. Our study was an endeavour to determine the feasibility of implementing a behaviour Change Communication Interaction developed for improvement in utilisation of maternity care services through male participation among the Saharia Tribes in Gwalior District, Madhya Pradesh.

**Methods:** The Study utilised a qualitative approach. Various activities were organised as a part of BCC, viz, Community mobilization, Campaign/Rallies, Interpersonal Communication-Drama & Mock Sessions, Face to Face counselling and Quiz etc. Action technique called Transect was used in order to know more about the environment and living of the people of Saharia Tribes in Gwalior District, Madhya Pradesh. Feasibility of the model was assessed by focusing on three main principles i.e., acceptability, integration and limited-efficacy testing.

**Result:** Acceptability testing study reveal that BCC intervention was successfully accepted by intended individual-both targeted individuals and those involved in implementing programs. Integration approach reveal that no major change in infrastructure of Govt. Programmes and facilities is required; only the participation of key people in society and workers local health services is required. Limited-efficacy testing reveals that there is a behavioural change in men’s perception of accompanying their spouse to the health centre; same was observed on the vaccination day in the village.

**Conclusions:** The BCC intervention proved to be feasible to implement. The Proposed BCC interaction is feasible and accepted by both Programme stake holders and beneficiaries.

## Background

The Indian tribal population is diverse, with a wide range of customs, ways of life, and cultural practices. However, there is one thing that all Indian tribal communities have in common: they have worse health indicators, a higher rate of illness and mortality, and very restricted access to medical care. Their health issues require extra consideration in the right setting. The participation of men in maternal health services is crucial for the overall growth of both mothers and their children. Many research findings show that when men are actively involved in maternal health, it strengthens the relationship within the family and also lowers the costs associated with health problems. The men who enthusiastically participate in reproductive activities are well informed about the govt facilities, vaccination, post and prenatal care during pregnancy which improves the utilisation of these services by the pregnant women and further curtail down the rate of maternal mortality. Studies have also indicated that couples who have good communication about pregnancy are more likely to raise their children in a healthier environment. [1,3]

It has been revealed in various studies that men participation in maternal health is rarely found, reasons associated with it are lack of knowledge of RCH services, illiteracy, addiction of alcohol, financial crisis, family matters, loss of wage, health centre resource constraints, Behaviour of health centre workers, lack of interest of policy makers, male dominance, attitude of elders in the home, laziness, personal attitude towards antenatal services, lack of trust on health workers, domestic violence and various others. Considering these obstacles, numerous studies highlight the necessity for creating more effective interventions and feasibility studies to overcome these barriers and to enhance men’s participation in maternal health services. [4]

One significant indicator to assess the status of healthcare in any nation is the maternal mortality ratio, or MMR. In the past 20 years, India has significantly decreased the number of maternal fatalities. The Maternal Mortality Ratio (MMR) in India was notably high in 1990, with 600 deaths per 100,000 live births, or nearly 1.5 lakh deaths annually. According to the most recent estimates from Sample registration system(SRS), under-5 mortality in India is 36/1000 live births(SRS,2018) and Neonatal Mortality Rate is 23/1000 live births(SRS,2018). MMR in the country has declined from 130 in 2014-15 to 113 in 2016-18; From past 27 years there was a 45% decrease on a global scale. To facilitate the worthy health care to pregnant women’s, various Programmes has been initiated by Govt. of India like Janani Suraksha Yojana (JSY), Janani Shishu Suraksha Karyakaram(JSSK), Pradhan Mantri Surakshit Matritva Abhiyan(PMSMA), Surakshit Matritva Aashwasan (SUMAN) etc. [5].

RCH services aims at combating and reducing the mortality rates of infant, mother and children. Numerous studies on Utilization of Health Services and RCH Services has been done which clearly depicts that districts that facilitate more RCH services also have better reproductive and child health. Therefore growth in the utilization of reproductive and maternal health services will not only curtail down the reproductive morbidities, but it will also reduce the child mortality [2]. Implementation of interventions in the concept of good health and disease prevention found to be both efficacious and effective. Due to resource constraints all interventions cannot be tested for both efficacy and effectiveness and hence, feasibility studies play a profound role in determining whether an intervention should be recommended for efficacy testing. To make favourable health behaviour changes, Behaviour change communication (BCC) is used to provide revive messages and a supportive environment that encourages individuals and communities to enhance their knowledge about health. [6].

BCC intervention plays a vital role in influencing the people’s social, environmental and organizational conditions as well as their preferences, notions and customs. However there are many empirical evidences which reveal that BCC intervention is of immense importance in bringing changes in the behaviour of community. To curtail down the child morbidity and mortality in poor communities Bangladesh Rural Advancement Committee (BRAC) initiated Improving Maternal neonatal and child survival (IMNCS) programme. The IMNCS programme introduced BCC interventions from its initiation in 2006. The motive of this study was to explore community perceptions of BCC interventions of the BRAC IMNCS programme in rural Bangladesh. BRAC is one of the largest NGOs in the world. The study revealed that the IMNCS BCC interventions had influenced both men and women to take health promoting decisions and seek maternal, neonatal and child health services. Remarkable changes such as improvement in healthy cord care practice, delayed bathing of the new-born and reduction of infant mortality was also observed in intervention districts as compared to that of comparison districts.[7,8,9]

A study on community perspective on men’s role in the utilization of maternal health care services among Saharia Tribes reveals that men seldom accompanied women for antenatal check-ups due to lack of knowledge, awareness, norms, beliefs, gender inequitable attitudes etc. There is a need for development of gender sensitive interventions that address the individual, community health care facility level barriers of male involvement. [10]. Saharias, one of the primitive tribes, are mainly located in the Chambal division, i.e., Gwalior, Morena, Guna and Shivpuri. Their total population is over 2 lac and is one of the poorest primitive tribe of Madhya Pradesh [11]. The main residential area of Saharia tribe is the forest of Shahabad which is spread from Rajasthan to Guna of Madhya Pradesh. Main source of income of Saharia Tribes is from agricultural work. They are generally influenced by Hindu culture; they worship many Hindu God and Goddesses and also celebrate various Hindu festivals. Our study was an endeavour to determine the feasibility of implementing a Behaviour Change Communication interaction developed for improvement in utilisation of Maternity care services through male participation among the Saharia Tribes in Gwalior District, Madhya Pradesh.

### Aims and objectives

The aim of the study was to determine the feasibility of implementing a behaviour Change Communication interaction for improvement in utilisation of Maternity care services through male participation among the Saharia Tribes in Gwalior District, Madhya Pradesh. The Maternity care services aims at providing at least three antenatal check-up, immunization against tetanus, and iron and folic acid for anaemia management.

### Methodology

There are number of assessment indicators for feasibility study but our study includes Acceptability, Integration and Limited-efficacy testing as described below.[12]

#### Acceptability

This relatively common focus looks at how the intended individual recipients—both targeted individuals and those involved in implementing programs—react to the intervention.

#### Integration

This focus assesses the level of system change needed to integrate a new program or process into an existing infrastructure or program. The documentation of change that occurs within the organizational setting or the social/physical environment as a direct result of integrating the new program can help to determine if the new venture is truly feasible.

#### Limited-efficacy testing

Many feasibility studies are designed to test an intervention in a limited way. Such tests may be conducted in a convenience sample, with intermediate rather than final outcomes, with shorter follow-up periods, or with limited statistical power.

### Study Design

The Study utilised a qualitative approach. BCC activities were planned initially to begin in one of the village for testing the feasibility of the model. Twenty male members from the village were selected for awareness training initially to engage them in the activities planned by the team and technical expert of BCC. The intervention was delivered for a period of one month. Pre-post awareness questionnaire was filled by the registered men. To check whether the BCC model might be integrated into the existing project few satisfaction interviews were also conducted among the programme delivery personnel.

### Participants

For the awareness training on RCH for Males, all men of 18 years or above age, who voluntarily agreed to participate, were enrolled for the 2 day training awareness programme.

### Procedure

Various activities were organised as a part of BCC like Community mobilization, Campaign/Rallies, Interpersonal Communication-Drama & Mock Sessions, Face to Face counselling and Quiz etc. Action technique called Transect was used in order to know more about the environment and living of the people of Saharia Tribes in Gwalior District, Madhya Pradesh.

### Process Prior to implementation of the BCC Model

To know more about the environment and living, a participatory learning & action technique called Transect was used. Transect is an observatory walk through the residential area of a village, observing and making notes of the layout of the village, housing, drainage, backyards, infrastructure, (schools, shops, wells, electricity), etc. It helps to locate/assess, map, and analyze various aspects of the residential area of the village that normally go unnoticed. A village transect is different from the other transects as it has greater focus on the dwelling zone of the village. It also provides extensive coverage of the entire household and economic activities which would probably be otherwise go unnoticed.

To explore more of local male tribes of the village they were called to assemble in a place where two-way communication could be held. A meeting was organized with all households at the Anganwadi to explain them the objective of the intervention in the area. Since it was open forum, opportunity was given to all male members and their female partners too to open up and share the views about existing health services available to them. Based on the information generated through the study and interim analysis, BCC intervention model was developed in consultation with the BCC expert.

### Training of trainers

The social workers employees under the study were given 4 days vigorous training on development of BCC and doing feasibility study under the able guidance of BCC expert from National Institute for Public Cooperation and Child Development, New Delhi.

### Implementation of the BCC Interaction for Limited-efficacy testing

BCC strategy was worked out in one of the tribal District of Gwalior. As per the baseline data, the BCC activities were planned initially to begin in one of the village. Activities like Awareness Training on RCH for Males, Community Mobilization, Campaign/Rallies, Interpersonal Communication, Face to Face counselling and quiz etc were organized as a part of BCC.

After a good discussion, it was mutually and participatively decided to choose 20 male members from the village for their awareness training in the beginning to engage them in the activities planned by the team and the Technical Expert of BCC. BCC must play a role in awakening the community response to ensure community-based health services critical to maternal and new born health are available. The awareness training programme for the male members of Saharia tribes was conducted at village in Dabra Block with the following agenda :-

**Table 1:** Awareness Training Programme.

|  |  |
| --- | --- |
| **Day 1** |  |
| Introduction about the programme | BCC expert |
| Pre-test<br>(Regarding knowledge of RCH facilities<br>(ANC/PNC/Delivery and Immunization) | Social worker under the project, ASHA,<br>Anganwadi Worker |
| Discussion About Maternal and Child Health<br>Facilities, ANC/PNC Delivery and Immunization | Child Development Public Officer<br>(ICDS) |
| Role & Responsibilities of Husband towards RCH<br>services Utilization | BCC expert + |
| Quiz and Prize Distribution |  |
| **Day 2** |  |
| Visit of Nearest health Centre | Could not be conducted due to strike |
| Discussion on Knowledge of Maternal and Child<br>Health facilities provided by government.<br>Barriers and Solutions for Male Participation | BCC expert, Social worker under the<br>project and CDPO |
| Post test<br>( Regarding knowledge of RCH facilities<br>(ANC/PNC/Delivery and Immunization) | Social workers and CDPO |

### Community Mobilization

Community gathering with registered eligible couples as well as other senior village members, local community health care providers and field employee’s .under the project was held in the village. BCC activities to enhance men’s participation in maternal and child health were discussed The male and females presented themselves in the course of the meeting. They provided their perspective on the BCC intervention activities conducted by field staff, local health workers and the BCC expert from NIPCCD. The impact of BCC activity was visible in their views on the role of men in maternal and infant health. Men in the village expressed more clearly their willingness to participate in maternal and child health services provided. The elderly people in the village also supported male involvement.

Drama performance was also conducted by the male members and the local community health care providers with the aim to enhance male participation in RCH services, viz, ANC, PNC and immunisation. House-to-house visits were also undertaken to gather individual perspectives and concerns about improving men’s participation in the RCH.

### Campaigns/ Rallies

An awareness generation on RCH and the involvement of men was planned and organized in the region. Many people were involved in the campaign, such as Village leaders, AWW helpers, School Children, ANM, ICDS Supervisors, training team of BCC, Youth club members and SHGs along with school going adolescent girls. Campaign target areas were Maternal Health & Nutrition, Family planning, Child Health & Nutrition, Routine immunization, Hygiene & Sanitation, adolescent health & Nutrition and reduction of Alcohol consumption. A rally was arranged with all of the members present, and the volunteers went around the town delivering the messages that had been prepared in advance. Video coverage of campaign was done.

### Modification

As per agenda a visit to nearest health centre and meeting with all the stake holders to discuss the BCC activities was planned but due to strike in the health centre on the scheduled day and non-availability of the stake holders this component could not be completed. As a result, we eliminated this element from our BCC interaction, and meetings were held with stakeholders based on their availability rather than necessarily with all of them at once.

### Wage loss compensation and Rewards for the active participants in the BCC programme

The male members registered in the village received compensation for lost wages for all the days spent in the BCC team. The male members of the village who supported the cause were given additional rewards. A total of rewards are divided into two categories, one is for active husbands, and the other is for active participation in the Drama. Five male members were selected from each category and rewarded in the form of backpacks and blankets. Awarded male members and two females (an elderly and an eligible female were interviewed and their views on BCC activities and male participation in MCH service facilities were taken.

### Ethic Approval

Ethic approval was gained from Institutional Ethics Committee vide Ref No. NIMS/Ethics Committee/2014 dated: May 15, 2014 for the overall study covering the feasibility study also. Written consent was taken from all the participants who participated in the main study, in the development phase. In the feasibility study there was no need of taking the consent.

## Results

The results are related to the three main areas of the feasibility study: limited effectiveness testing, acceptance, and integration.

### The Participant flow diagram of the study

**Fig 1.**
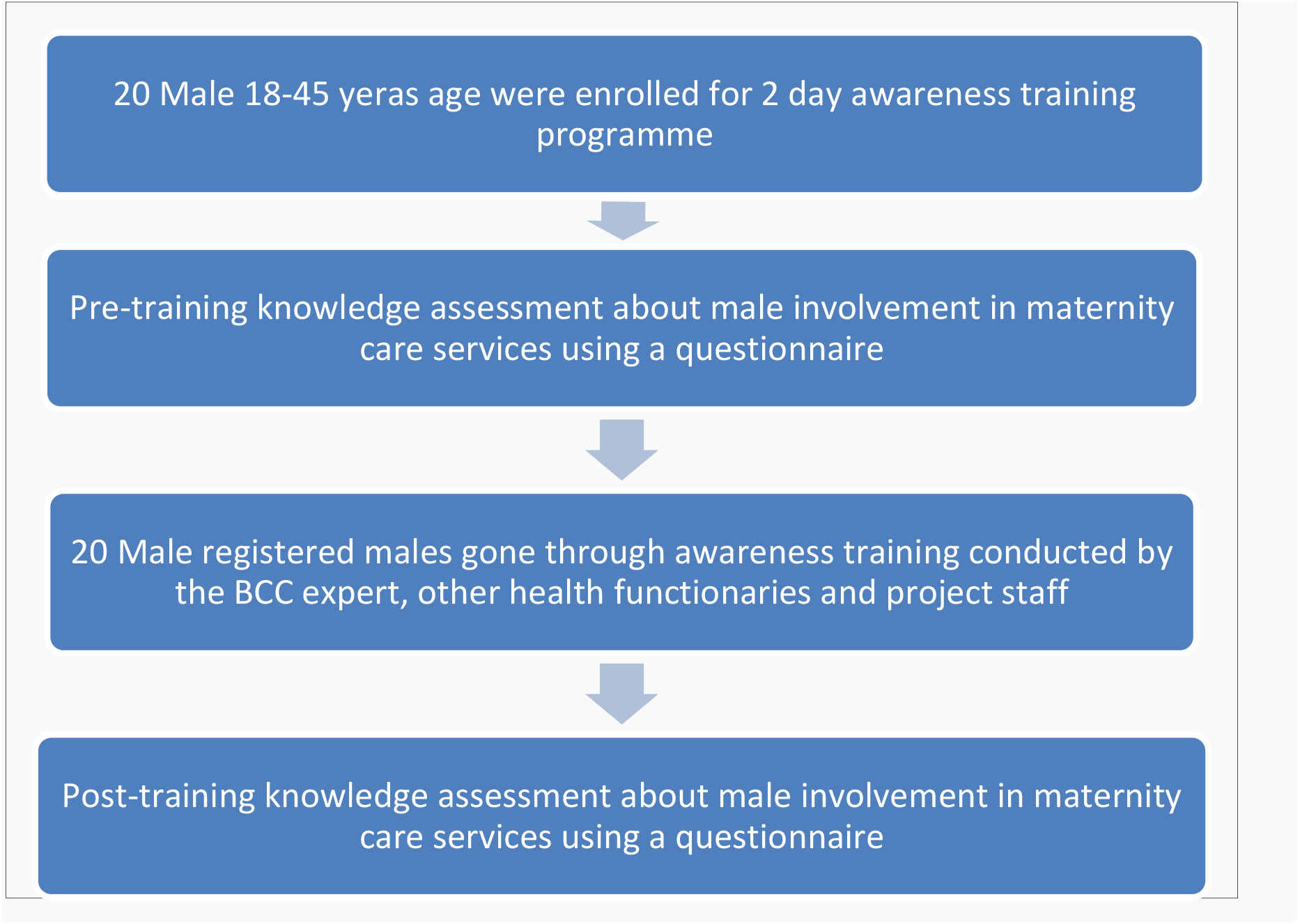
Consort Diagram of the Flow of participants of the 2 day training awareness programme.

### Acceptability

#### Targeted individual’s reaction to the intervention

Community gathering with registered eligible couples as well as other senior village members, local community health care providers and field employee’s under the project was held in the village. The male and females presented themselves in the course of the meeting. Men seem to be more vocal in expressing their willingness to take part in the maternal and child health services being provided by the govt. The village’s elders also supported male involvement. Couples appeared to be passionate about the entire idea. They had been of the view that such forms of activities were by no means being performed before, with-in the area and were very much optimistic about impact of BCC activity. But they had been worried concerning the bad behaviour the various male viz alcoholism, which of their opinion is going to be one of the boundaries in enhancing the male involvement in MCH services.

Couples enthusiastically expressed their views on the BCC intervention activities being carried out by the field staff, local health workers and expert from NIPCCD. They were of the view that these kinds of activities will enhance the role of men in maternal health care and will bring fruitful consequences in favour of women.

#### Reaction of Individuals’ involved in implementing programs

The individuals (health providers) involved in the implementation of the BCC model programme, also some of the key informants, were asked about their views on the activities under the BCC programme using a feedback form. The respondents were asked about the benefits and process of implementation of the activities under the proposed BCC model programme.

The respondent believed that the proposed knowledge and awareness program would lead to greater awareness of government programs and health facilities provided under the programs. It would change the behaviour of society and would also improve male participation and the health of both the mother and the child in general. It would also reduce maternal and infant morbidity and mortality in society. The awareness program would also improve mutual trust and coordination between couples in the community.

#### Feedback on community level activities under the BCC model

The respondent believed that the activities proposed at the community level would improve the attitudes of individuals and the community at large towards the husband’s involvement in maternal and child health activities. The proposed activities would create an environment conducive to the participation of men in maternal and child health and the male members of the village would no longer be ashamed to accompany their wives to the hospital for ANC / PNC, vaccination and delivery. This would improve the proportion of institutional deliveries and have a better impact on maternal and child health. The Nukkad Games of the BCC program would support the process of improving male participation in maternal and child health.

#### Feedback on quiz conducted with registered men

A quiz was conducted before and after the intervention of the BCC Model with the help of stakeholder groups and ANW. Various questions like where the delivery of pregnant women should be done? Did Husband should accompany his wife to health centre? Is it Mandatory to vaccinate new born and expected mothers? Do pregnant women need a nutrition regime as opposed to a normal diet? Did u felt ashamed of accompanying pregnant women to health centre? If you did not accompany your wife to health centre, did she able to get more beneficial health services? If you did not accompany to health centre will u able to fulfil your responsibility as a husband? Will you give time to your wife and new born after delivery?

#### Outcome of the Quiz

Drastic change was seen after the intervention. 100% of the men agreed that delivery should take place in a health centre, that the husband should accompany his wife to the health centre, that both the new born and the expectant mother must be vaccinated, and that they understood the importance of a proper nutritious diet and accompanying their wife to the health centre. Following the intervention, 87.5% of men stated that they would try to accompany their wife for delivery even if they were engaged somewhere else. Before the intervention, only 62.50% of men were of the opinion that they would accompany their wives to health centre after intervention 87.50% men agreed for the same. Outcome of the pre and post intervention is depicted in Figure 2.

**Figure 2:**
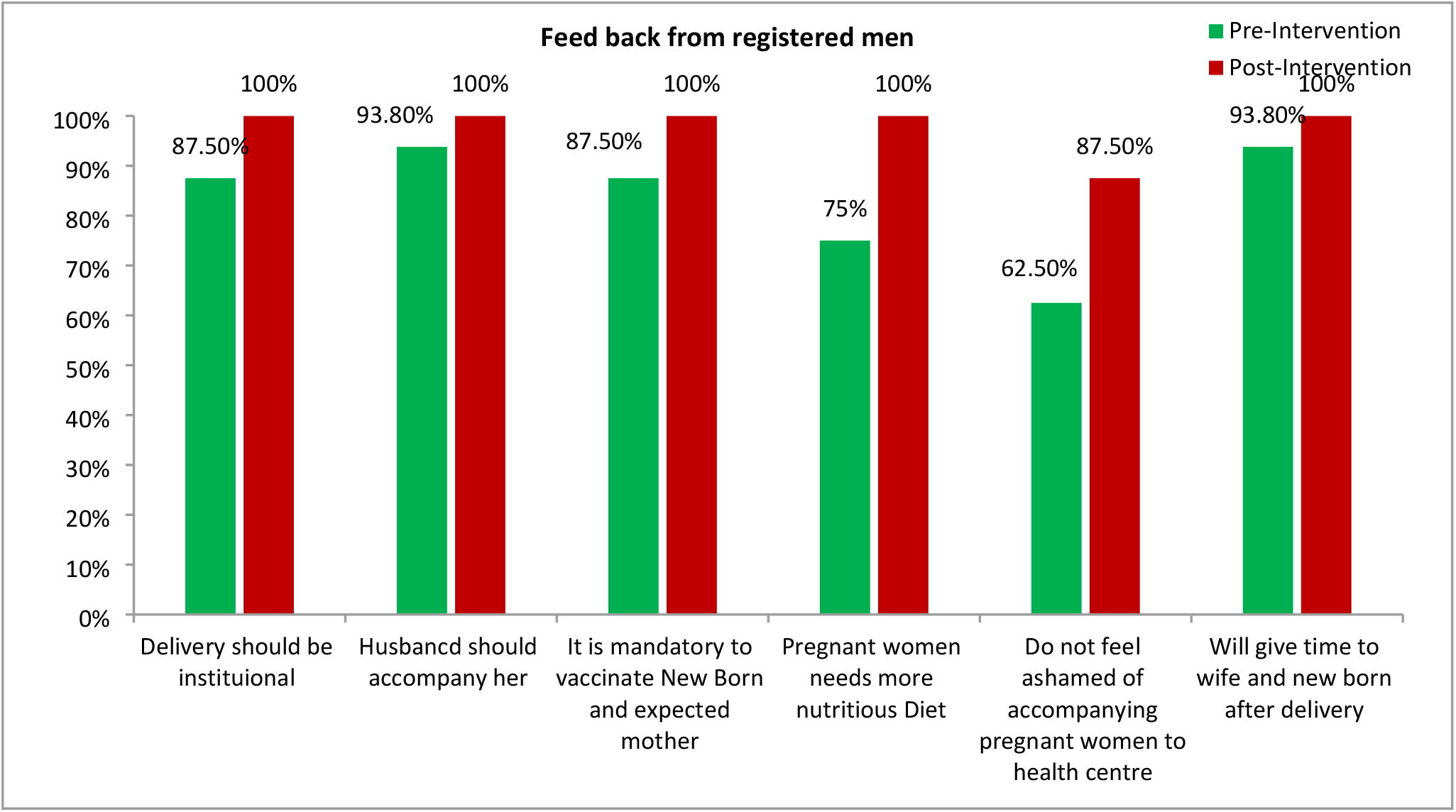
Short term impact of the BCC intervention on the knowledge and perception of registered men.

#### Feedback on home visits for couple counselling

The respondent believed that the proposed home visits for couple counselling would improve the knowledge of the families in the society about the benefits of the male participation in maternal and child health. Through this process the problems and believes of couples could be handled individually; this would help in accelerating the process of male participation.

### Integration

This approach assesses the degree of system change required to integrate the BCC model program with the existing infrastructure. And according to the components of our BCC model, the required system change is minimal. The various components of the Model BCC require different system changes.

1. Based on the awareness rallies and Nukkad Nataks (Street Plays) that are part of the community participation in the behavior change communication model, no major infrastructure change is needed, only the participation of key people in society and workers local health services is required.
2. The awareness program for the of eligible men and women group meetings, and the home visits of local health workers requires an additional component, namely, the participation of men in maternal and child health in the duties of the local health workers and improved level of commitment from them for success of the BCC model.
3. The component that requires the greatest degree of change in the existing system is not the BCC model component, but the required effect of the BCC, i.e. the male participation itself. In the existing system, in most of the gynaecological departments, men (any relatives) are not allowed to enter the ANC clinic. For this impact to take place, the support of state level health delivery system level stake holders is a must and needs rigorous effort to be made.

### Limited effectiveness testing

The short-term effect of the BCC intervention carried out in one of the Village was visible in registered couples as part of the awareness program. The projected intervention is implemented in the area for a short period of time, say one month, and the BCC intervention short-term impact was evaluated after one month. To assess impact, a community meeting of project researchers was held in the village with registered eligible couples along with other older village members, local health care providers and field workers as part of the project. Some of the villagers were also interviewed during the short-term impact assessment.

1. Villagers believed that male members of society should be actively involved in the use of maternal and child health services, namely prenatal care, maternal delivery and vaccination. Male members must accompany their wives to prenatal care visits, vaccination visits, and hospital deliveries. They must ensure that the pregnant woman takes 100 IFA tablets provided by the health center.
2. The practice in the past has been to give birth at home and for institutional deliveries and for ANC / PNC visits, generally the mother-in-law or some elderly women accompanied the woman to the health center. But after the awareness program and other BCC activities in the village, their mind changed and most of the people were in favor of her husband accompanying the woman to the health center. They believed that the amount of logistical support that a husband can provide to the wife cannot be provided by anyone else, including the elderly woman in the home.
3. During this brief period, there was a visible change in men’s perception of accompanying their spouse to the health center. A great change was observed in the presence of men on the day of vaccination.

## Discussion

The aim of the study was to determine the feasibility of implementing a behaviour Change Communication interaction for improvement in utilisation of Maternity care services through male participation among the Saharia Tribes in Gwalior District, Madhya Pradesh. BCC activities were planned initially to begin in one of the village for testing the feasibility of the model. The study explored three major areas of the feasibility study i.e. limited efficacy testing, acceptability and integration.

From the Acceptability testing, study reveal that by implementing the BCC interaction people became more aware of facilities provided by the Govt. Couples seems to be more enthusiastic about the entire idea. They were of the view that such kinds of activities were never conducted in their area. But they had been worried concerning the bad behaviour the various male viz alcoholism, which of their opinion is going to be one of the boundaries in enhancing the male involvement in maternity care services. The respondents were of the view that due to intervention mortality rate among women will decline and men will no longer felt ashamed to accompany their wives to health centre.

Integration approach reveals that no major infrastructure change is needed; only the participation of key people in society and workers local health services is required. For this impact to take place, the support of state level health delivery system level stake holders is a must and needs rigorous effort to be made. Short term impact of the BCC intervention shows that there is a behavioral change in men’s perception of accompanying their spouse to the health center. A great change was observed in the presence of men on the day of vaccination. These results are in line with the results of earlier studies which reveal that behavioural interventions have fruitful results. A study on self –compassion is a proof-of-concept study of an online adapted mindfulness-based compassionate living program, which reveals fruitful results patients who were earlier suffering from harsh self-criticism after intervention self-compassion, reassuring-self and satisfaction with life significantly increased whereas inadequate self, hated self, perceived stress and fear off self-compassion decreased.[13]

Various studies on tribal population reveals the same fact that alcoholism is identified to be one of the biggest impediments in men’s involvement in maternal health care.[10] A study on mortality among Saharia reveals that poor eating habits, negligence of iron, calcium, vitamins and maternal care during pregnancy from the inception of the pregnancy to its termination are all factors that contribute to maternal mortality among Saharia women [14].Our study, which used the BCC model, resulted in a variety of changes in the behaviour of elderly people, both men and women, as well as a shift in their attitudes on men’s engagement in maternal care. People become more knowledgeable about the government’s numerous services.

Moreover, health status of tribal women in India their access to employment and men’s role in the utilization of maternal health care services among Saharia Tribes reveals that there is a need for development of gender sensitive interventions that address the individual, community health care facility level barriers of male involvement.[15,10] Our study, which used the BCC model, focused on increasing male engagement in utilization of maternity care services. Men who were previously hesitant to accompany their wives to the health centre now understand the value of ANC and PNC visits after participating in the intervention programme.

### Limitations

In the proposed BCC interaction there was a provision to visit nearest health centre but due to strike on the scheduled day of visit we are unable to fulfil the task hence this element was eliminated from the study. In the proposed BCC interaction it was planned that co-workers will supposed to conduct a meeting with all the stake holders to discuss the BCC activities but due to non-availability of the stake holders this component was modified and meetings were held with stakeholders based on their availability rather than necessarily with all of them at once.

As the BCC intervention is a customized communication package for a specific group and in our study it is customized and developed for Tribal population of Saharia tribes, Madhya Pradesh. The proposed BCC interaction feasibility study was customized for two blocks of Gwalior district Madhya Pradesh. Hence it can be generalized for the whole tribal community of Madhya Pradesh state only.

### Future endeavours

After testing the feasibility of the Proposed BCC intervention, we have initiated a community trial for assessing the impact of the BCC on male involvement and utilisation of maternal care services in four districts of Madhya Pradesh.

## Conclusion

Motive of our study was to determine the feasibility of implementing a behaviour Change Communication interaction for increasing male engagement in utilisation of maternity care services among the Saharia Tribes in Gwalior District, Madhya Pradesh.

The BCC intervention proved to be quite fruitful, as changes in the behaviour of both men and women as well as the elderly in the hamlet, were noted. Men no longer hesitate to accompany their wives to ANC, Institutional delivery and PNC visits after intervention. They become more aware of the facilities provided by the Govt. No major changes to the infrastructure are required, just the participation of key people in society and services of local health workers is required. Those who participated in the implementation of the program believed that it would also reduce maternal and infant morbidity and mortality in society. In a nutshell, this type of study can significantly change people’s lives/attitudes. The Proposed BCC interaction is feasible and accepted by both Programme stake holders and beneficiaries.

## Data Availability

Data will be made available

NA

## Abbreviations

BCC: Behaviour change communication
RCH: Reproductive and child health
BRAC: Bangladesh Rural Advancement Committee
ANC: Antenatal care
PNC: Postnatal care
IMNCS: Initiated Improving Maternal neonatal and child survival.

## Supplementary Information

### Authors’ Contributions

Conceptualisation of the study : TA, SN, Development of Methodology: TA, SN, Analysis: TA,SN, AK; Drafting of the manuscript: TA,SN, AK Critical Review: TA, SN

### Funding

The project was funded by Indian Council of Medical Research. However the project was completed, the report has been submitted and there are no more funds available for the research paper to be published.

## References

1. Salil Basu. Dimensions of tribal health in India, Health and Population Perspectives and Issues23(2): 61–70, 2000.

2. Sharma RK, Ranjan R, Kumar A, Pandey A. Utilization of health services and RCH status in Madhya Pradesh: a District Level Analysis. In Proceedings of National Symposium on Tribal Health 2011.

3. Bagenda F, Batwala V, Orach CG, Nabiwemba E, Atuyambe L. Benefits of and barriers to male involvement in maternal health care in Ibanda District, southwestern, Uganda. Open Journal of Preventive Medicine. 2021 Dec 21;11(12):411–24.

4. Davis J, Vyankandondera J, Luchters S, Simon D, Holmes W. Male involvement in reproductive, maternal and child health: a qualitative study of policymaker and practitioner perspectives in the Pacific. Reproductive health. 2016 Dec;13:1–1.

5. Department of Health & Family welfare, Ministry of Health & Family Welfare, Government of India. Annual Report. 2021-22.1-464p.Available from https://www.mohfw.nic.in.[Accessed 22 March 2024].

6. Ciara Briscoe, Frances Ab345345-oud. Behavior change communication targeting four health behaviors in developing countries: A review of change techniques. Social Science & Medicine 75(2012) 612–621.

7. Salam SS, Khan MA, Salahuddin S, Choudhury N, Nicholls P, Nasreen HE. Maternal, neonatal and child health in selected Northern Districts of Bangladesh: findings from baseline survey 2008. Dhaka: BRAC Reports; 2009.

8. Rahman, A., Leppard, M., Rashid, S. et al. Community perceptions of behaviour change communication interventions of the maternal neonatal and child health programme in rural Bangladesh: an exploratory study. BMC Health Serv Res 16, 389 (2016). 10.1186/s12913-016-1632-y.

9. Rahman M, Jhohura FT, Mistry SK, Chowdhury TR, Ishaque T, Shah R, et al. (2015) Assessing Community Based Improved Maternal Neonatal Child Survival (IMNCS) Program in Rural Bangladesh. PLoS ONE 10(9): e0136898. 10.1371/journal.pone.0136898

10. Sarita Nair, Tulsi Adhikari, Atul Juneja, Bal Kishan Gulati, Ashpinder Kaur & M. Vishnu Vardhana Rao. Community perspectives on Men’s Role in the Utilisation of Maternal Health Services Among Saharia Tribes in Gwalior, Madhya Pradesh, Inida: Insights from a Qualitative Study. Maternal and Child Health Journal. DOI 10.1007/s10995-020-03029-8.

11. Tapas Chakma, P. Vinay Rao, P.K. Meshram, S.B. Singh. Health and Nutrition Profile of Tribals of Madhya Pradesh and Chhattisgarh Proceeding of National Symposium on Tribal Health(2006).

12. Bowen DJ, Kreuter M,Spring B, etal. How We Design Feasibility Studies. Am J Prev Med. 2009;36(5):452–7.10.1016/j.amepre.2009.02.002.

13. Tobias Krieger, Dominik Sander Martig, Erik Van den Brink, Thomas Berger. Working on Self-Compassion Online: A proof of Concept and feasibility study. Internet interventions 6(2016) 64–70. 10.1016/j.invent.2016.10.001.

14. Ranjan Kumar Biswas & A.K. Kapoor (2003) A study on Mortality among Saharia-A Primitive Tribe of Madhya Pradesh, The Anthropologist, 5:4, 283–290, 10.1080/09720073.2003.11890820.

15. Basu SK (1993). Health Status of Tribal Women in India. Social Change 23(4). P 19–39.

